# Evaluating the impact of demographic, socioeconomic factors, and risk aversion on mobility during COVID-19 epidemic in France under lockdown: a population-based study

**DOI:** 10.1101/2020.05.29.20097097

**Authors:** Giulia Pullano, Eugenio Valdano, Nicola Scarpa, Stefania Rubrichi, Vittoria Colizza

## Abstract

On March 17, 2020, French authorities implemented a nationwide lockdown to respond to COVID-19 epidemic and curb the surge of patients requiring critical care, similarly to other countries. Evaluating the impact of lockdown on population mobility is essential to quantify achievable reductions and identify the factors driving the changes in social dynamics that affected viral diffusion. We used temporally resolved travel flows among 1,436 administrative areas of mainland France reconstructed from mobile phone trajectories. We measured mobility changes before and during lockdown at both local and country scales. Lockdown caused a 65% reduction in countrywide number of displacements, and was particularly effective in reducing work-related short-range mobility, especially during rush hours, and recreational trips. Geographical heterogeneities showed anomalous increases in long-range movements even before lockdown announcement that were tightly localized in space. During lockdown, mobility drops were unevenly distributed across regions. They were strongly associated with active population, workers employed in sectors highly impacted by lockdown, and number of hospitalizations per region, and moderately associated with socio-economic level of the region. Major cities largely shrank their pattern of connectivity, reducing it mainly to short-range commuting. Lockdown was effective in reducing population mobility across scales. Caution should be taken in the timing of policy announcements and implementation, as anomalous mobility followed policy announcements that may act as seeding events. On the other hand, risk aversion may be beneficial in further decreasing mobility in largely affected regions. Socio-economic and demographic constraints to the efficacy of restrictions were also identified. The unveiled links between geography, demography, and timing of the response to mobility restrictions may help design interventions that minimize invasiveness while contributing to the current epidemic response.

**Funding:** ANR projects EVALCOVID-19 (ANR-20-COVI-0007) and DATAREDUX (ANR-19-CE46-0008-03); EU H2020 grants RECOVER (H2020-101003589) and MOOD (H2020-874850); REACTing COVID-19 modeling grant.

## INTRODUCTION

French authorities responded to the rapid growth of COVID-19 cases by imposing heavy restrictions on mobility, as many other countries in Europe and beyond^1^. Lockdown was enforced on March 17, 2020, and helped slow down infection rates and limit the strain on the healthcare system^2^. As these restrictions are gradually being phased out, it is essential measure changes in human mobility to (i) quantitatively determine how imposed measures and recommendations (e.g. regarding telework where possible, ban of leisure trips) translated into reduced mobility at specific scales and times, (ii) inform models estimating the effectiveness of the ongoing lockdown in reducing the epidemic spread^2,3^, (iii) identify the driving factors associated to documented reductions to help devising social distancing measures needed for the post-lockdown phase. Accessing human mobility data is now possible at several spatial and time scales, and often in nearly real-time. These data have been proven useful in many epidemiological contexts^4^ – including for example the West Africa Ebola epidemic^5^ – and are being used now for COVID-19 pandemic in many countries^6^ – namely, Belgium^7^, Germany^8^, India^9^, Italy^10^, Poland^11^, Spain^12^, UK^13^, USA^14,15^.

Mobile phone records are one of the main sources of mobility data. They describe travel flows among the different locations of a country. These flows can be analyzed over time to study population patterns, with no information on individual users, safeguarding privacy^6^. In this study, we used data provided by Orange Business Service Flux Vision, and studied how mobility in France changed before and during lockdown. We broke down our results by trip distance, user age and residency, time of day, and analyzed regional data and spatial heterogeneities. We investigated behavioral responses to announcements of interventions, and to the epidemic burden, as well as associations of mobility reduction with demographic and socioeconomic indicators. Considering the network of travel connections among French locations, we also identified the most vulnerable and most resilient connections to the mobility shock induced by lockdown, with a specific focus on main French cities.

## METHODS

### Timeline of COVID-19 epidemic in France

Three phases have marked the French response to COVID-19 epidemic (**Figure 1**). Phase 1 started on January 10, 2020. Its aim was to detect imported cases and identify local transmissions through case-contact investigations. Phase 2 started upon appearance of localized clusters, and added targeted social distancing interventions around reported clusters (e.g. school closure, gatherings and public transport bans). Phase 3 was declared on March 14, 2020 when the virus was recognized to actively circulate in the territory.

**Figure 1.**
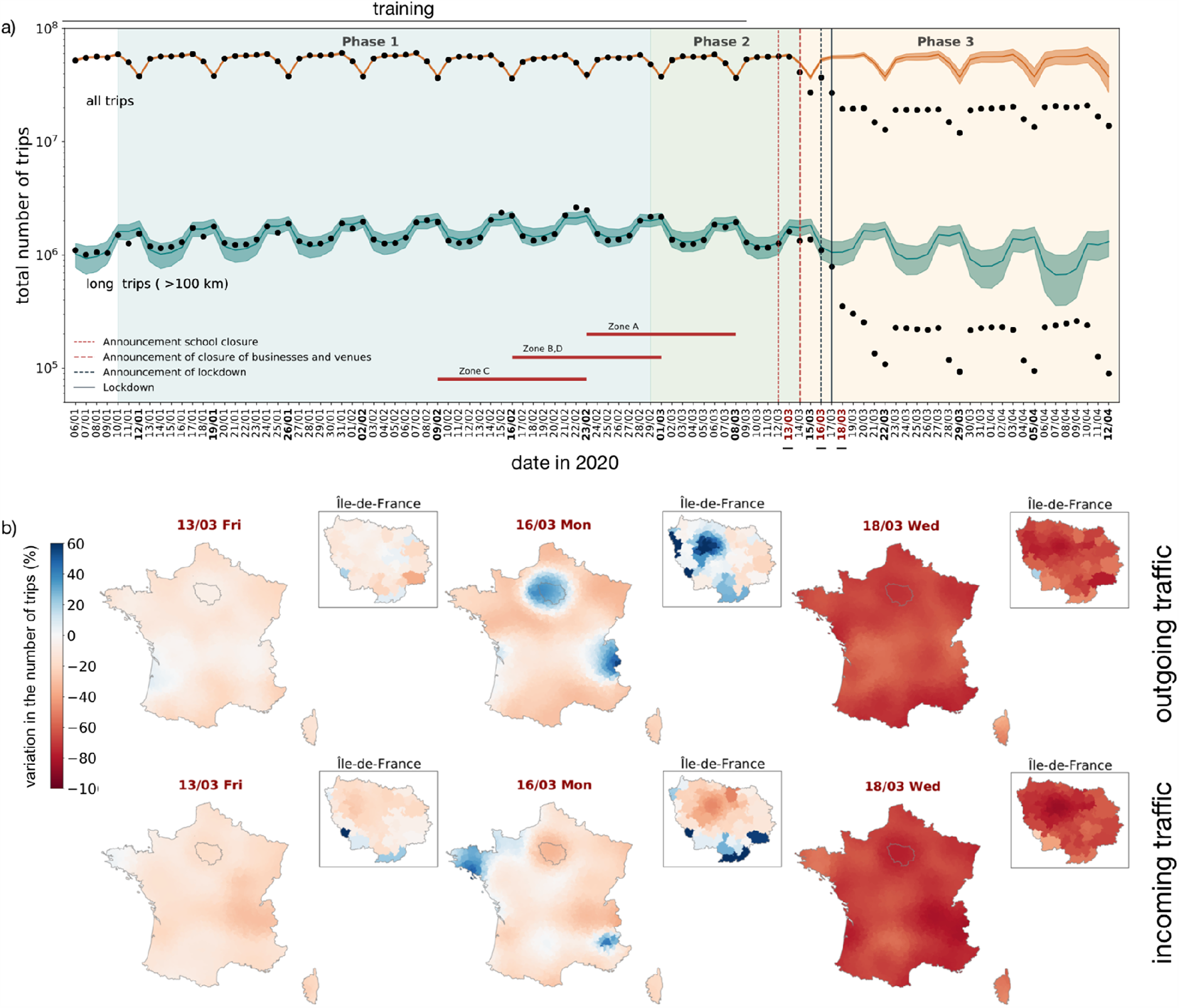
The phases of the COVID-19 epidemic in France, and its impact on mobility patterns. a) Colored areas correspond to the different phases of the epidemic response. Red lines mark main government interventions. Red horizontal thick lines indicate school holidays (students staying home from school) in the four government-defined geographic areas of France (Paris is in zone C). Dashed vertical lines indicate announcements made by French authorities. March 12: announcement of school closure to be implemented starting March 16; March 14: announcement of closure of nonessential businesses with immediate effect; March 16: announcement of lockdown to be implemented the day after at noon. The black solid vertical line on March 17, 2020 indicates the beginning of lockdown. The black dots track the temporal change of the total number of daily trips measured from mobile phone data in France from January 6 to April 12. The top timeline considers all trips, the bottom timeline only long trips (>100 km). Each timeline is fitted (orange – all trips, green – long trips), with the training set going from January 6 to March 9, and extrapolation up to April 12. A solid horizontal black line above the plots indicates the span of the training set. Shaded areas represent 95% confidence intervals. b) Maps show the variation in traffic compared with the unperturbed baseline predicted by the fit. Top row: outgoing traffic; bottom row: incoming traffic. The chosen dates are March 13, March 16 (day before lockdown), March 18 (day after lockdown enforcement). Ile-de-France is highlighted in the zoomed panel (the region is indicated by its contour in the map of France).

### Data

Mobility data were provided by the Orange Business Service Flux Vision in the form of displacement matrices. They comprised origin-destination travel flows among 1436 geographic areas of mainland France. For each pair of locations and any given day, data were provided stratified by age class (less than 18 years old, 18-64, 65 and more). Each area belongs to one of 13 regions, which are the subnational administrative divisions of mainland France. Details on data format and extraction in Appendix S1.

Regional hospitalization data were obtained from Santé publique France^16^.

We used population data and regional socioeconomic indicators from the French National Statistical Institute (INSEE)^17^: i) Fraction of population in the age range 24-59, which includes most of the population in working age^17^. ii) Standard of living – 9^th^ decile, defined as the 9^th^ decile of the household’s gross disposable income divided by the number of consumption units (which measure household size: one unit for the first adult, 0.5 units for each additional person over 14 years of age and 0.3 for each child under 14 years of age).

Employment data were obtained from INSEE, and the French Ministry of Labor^18^. As indicator, we used the fraction of employed workers in the sectors mostly affected by lockdown. These are the sectors in which at least 50% of employees stopped working (hotels, hospitality, food services, and construction), or had been working remotely (finance, insurances, IT)^18^.

### Ethics

Mobile phone data were previously anonymized in compliance to strict privacy requirements, presented to and audited by the French data protection authority (CNIL).

### Timeline fit and prediction

To fit and forecast time series we used the forecasting procedure Prophet by Facebook Open Source^19^. Prophet decomposes a time series into non-periodic and periodic components which then are fitted using MCMC (Appendix S2). We fitted Prophet on traffic flow data from January 6, 2020, to March 9, 2020 (training set of the model), and extrapolated traffic flow after March 9, assuming no perturbation due to COVID-19 or associated interventions. We then measured the deviation of observed traffic from the predicted temporal evolution of unperturbed traffic over time.

### Trip analysis

Our analyses were performed on all trips and on trips whose geodesic distance between location centroids is longer than 100 km (*long trips*). This cutoff effectively discards commuting, as ∼95% of daily work-related trips are shorter than 100 km^17^. We distinguished between residents, i.e., users with French SIM cards, and foreigners. We broke down data in the three age classes. We classified trips by their time of day: daytime (7am-7pm), nighttime (7pm-7am), and distinguished between weekdays and weekends. During weekdays we also considered rush hours (7am-9am, 5pm-7pm).

### Mobility reduction during lockdown

Computed in a case-crossover framework by comparing the week starting Monday April 6, 2020 (3 weeks into lockdown), to the week starting Monday February 3 (control week). The latter was chosen as being before school holidays, and after strikes of public transport.

### Statistical analysis

All statistical analyses were performed in R, version 3.6.1. Two-sided significance of Pearson coefficients was determined at a level of 0.05. The coefficient of variation is defined as the sample standard deviation divided by the sample mean, expressed in percentage points.

### Network analysis

Nodes in the networks represent the geographic locations in which we divided mainland France, and links represent trips between locations. Links are directed (trips have origins and destinations), weighted (by the number of trips linking two locations), and evolve in time. We used standard Python libraries, among which NetworkX. Link persistence probability at a given week was defined as the probability that a connection present in the network during week February 3-9 (benchmark week) was still present in the week under consideration.

### Maps

To smooth spatial data, we used a gaussian kernel (Appendix S3). The radius containing 95% of outgoing traffic from a city was computed by considering all mobility links that start from that city, each with its geodesic distance. They were included incrementally from the shortest to the longest, until the cumulative sum of the weights of the included links reached 95% of the total outgoing traffic.

### Role of the funding source

The funders had no role in study design, data collection, data analysis, data interpretation, writing of the manuscript, and decision to submit. All the authors had full access to all the data used in the study and had final responsibility for the decision to submit for publication.

## RESULTS

### Behavioral response during the transition period up to lockdown implementation

While no observable change in mobility occurred during Phase 1 and 2 of the epidemic, the start of Phase 3 on March 14 had a substantial impact on mobility in France (**Fig. 1a**). This transition occurred prior to the announcement (March 16) and implementation (March 17) of lockdown measures, and saw nationwide mobility go from ∼60M trips per day down to ∼20M trips after lockdown entered into effect. The shock in mobility spread out over a transition period lasting almost a week.

Starting March 14, 2020, total flow was significantly below the predicted unperturbed traffic (outside 95% credible interval), as a likely consequence of the start of Phase 3. Mobility further decreased on Sunday, March 15, when local elections took place. Instead, a rise in traffic took place on the day before lockdown enforcement. Traffic volume on that day was markedly higher than the surrounding days, but still lower than the predicted baseline, and possibly compatible with the typical Sunday-to-Monday pattern. Long trips (>100 km) were also significantly – albeit slightly - below the predicted baseline during the weekend (March 14, 15). They however went back to seemingly normal values on March 16 – i.e., in agreement with the unperturbed prediction -, and near-to-normal values on lockdown day. However, this country-level behavior hid anomalous deviations from the predicted mobility behavior in specific locations, as **Fig. 1b** shows. Spikes in outgoing traffic are distinctively visible in Île-de-France (the region of Paris) and, at the same time, in incoming traffic in Normandy and Bretagne. They measure the pre-lockdown exodus out of Paris occurring before lockdown took effect^20^. Analyses at finer scales within Île-de-France revealed that anomalous outgoing traffic concentrated in the Paris area, and western Île-de-France. Similar spikes of outgoing and incoming traffic were also visible in the South East, close to the Alps, as reported previously^20^.

### Mobility during lockdown

Mobility patterns quickly entered a new equilibrium after lockdown enforcement, marking the end of the transition period. Using a case-crossover framework (see Methods), we found that lockdown decreased the overall number of trips by 65% (20M over 57M trips per day) (**Figure 2a**). Reduction was stronger for trips made by foreigners (∼85%, 0.25M over 1.62M). Their number of trips was however very small even before lockdown compared to French residents (3%), therefore we excluded them from the rest of the analysis. Long-range traffic (>100 km) was disrupted more severely than average (85% reduction, 0.24M over 1.7M, **Fig. 2a**). This was likely associated with a disruption of long-range transportation (trains, flights), and the ban of leisure-related trips, also confirmed by the almost disappearance of long trips during the weekend (see below).

**Figure 2.**
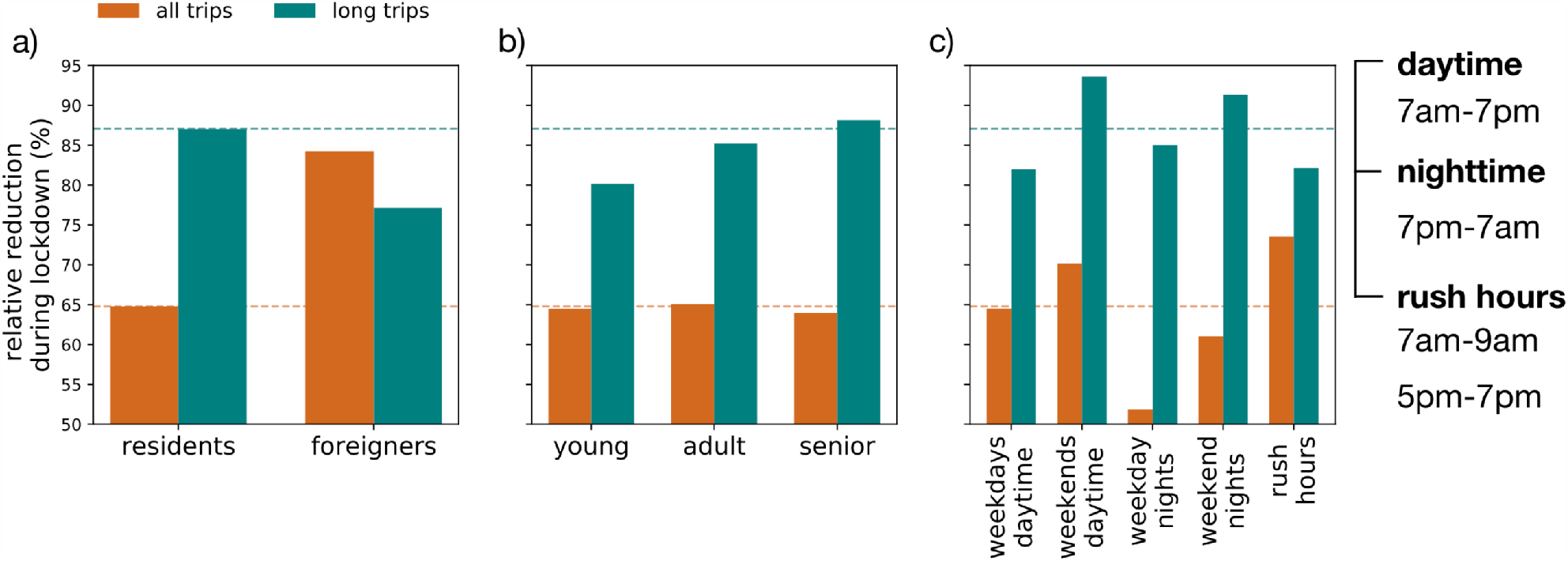
Mobility reduction during lockdown across user type, age and time of day. Reduction is computed as the average over the week starting Monday April 6, with respect to the average over the first week of February (starting Monday February 3). a), b) and c) show the relative reduction broken down by residents/foreigners, age classes, and times of the day. They also show statistics for all trips (orange) and long trips (green), defined as trips with geodesic distance longer than 100 km. Horizontal orange and green lines indicate relative reduction on all residents (all, long, respectively).

Mobility reduction in total trips was homogeneously distributed across age classes (**Fig. 2b**). When considering only long trips, reduction instead increased with age, as seniors reduced their trips above 100 km by ∼ 90% (30k over 250k).

Drops in mobility were uneven across the time of the day (**Fig. 2c**). Movements during rush hours were the most disrupted, the combined effect of school closure and telework leading to a ∼75% reduction (1.2M over 4.6M trips per hour). Daytime movements during weekends also exhibited a higher-than-average decrease, hinting at a successful reduction of recreational activities. Nighttime movements during weekdays instead recorded the lowest reduction, well below average. They might be related to unavoidable work-related mobility, whose impact is however likely to be limited, as these movements make up for only ¼ of the total. Long-range mobility almost completely stopped during weekends (around 95% decrease, 0.17M over 3M trips per day).

### Regional heterogeneities in mobility reduction during lockdown

Traffic reductions were not homogeneous across the 13 regions of mainland France. Reduction of internal traffic was above average in 4 regions (Île-de-France, Auvergne-Rhône-Alpes, Grand Est, Provence-Alpes-Côte d’Azur), whereas markedly below average in Bourgogne-Franche-Comté, Centre-Val de Loire, and Normandy (**Figure 3**). Similar fluctuations were visible in outgoing traffic (coefficient of variation equal to 8.4% compared to 8.0% for internal traffic). Île-de-France, Hauts-de-France and Grand Est all experienced above-average reductions in outgoing mobility, as high as 80% (117k over 585k) for Île-de-France. Corse also exhibited a reduction comparable to Île-de-France, showing a clear disruption of the long-range connections linking the island to mainland France. Similar reductions were obtained with incoming fluxes in the regions (not shown).

**Figure 3.**
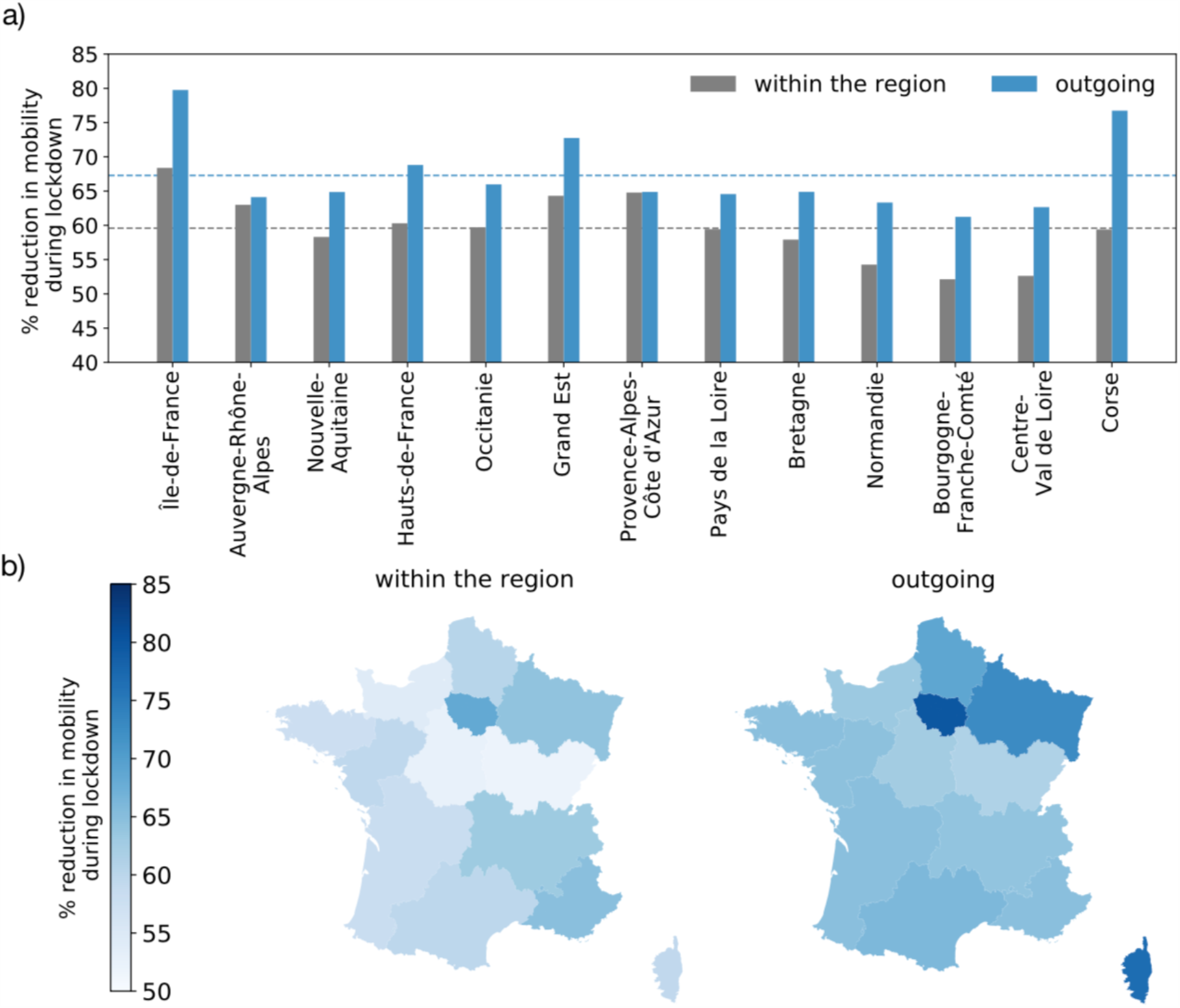
Lockdown-induced mobility reduction across regions. a) Breakdown of mobility reduction by region in mainland France. Reductions for trips within region are in gray, for trips leaving the region are in blue. The horizontal gray and blue lines indicate the corresponding averages across regions. b) Map visualization of a).

The impact of nationwide lockdown in the reduction of outgoing mobility per region was strongly associated with the fraction of the population in the most active age range (24-59 years old)^17^ (Pearson r = 0.91, p < 0.01) and the fraction of workers employed in sectors that substantially modified their organization during lockdown, due to telework, partial or full closure of activities (Pearson r = 0.80, p < 0.01) (**Table S1** and **Figure 4**). It was moderately associated with regional economic disparities, in terms of a positive correlation with the 9^th^ decile of the standard of living of the region (Pearson r = 0.63, p = 0.02).

**Figure 4.**
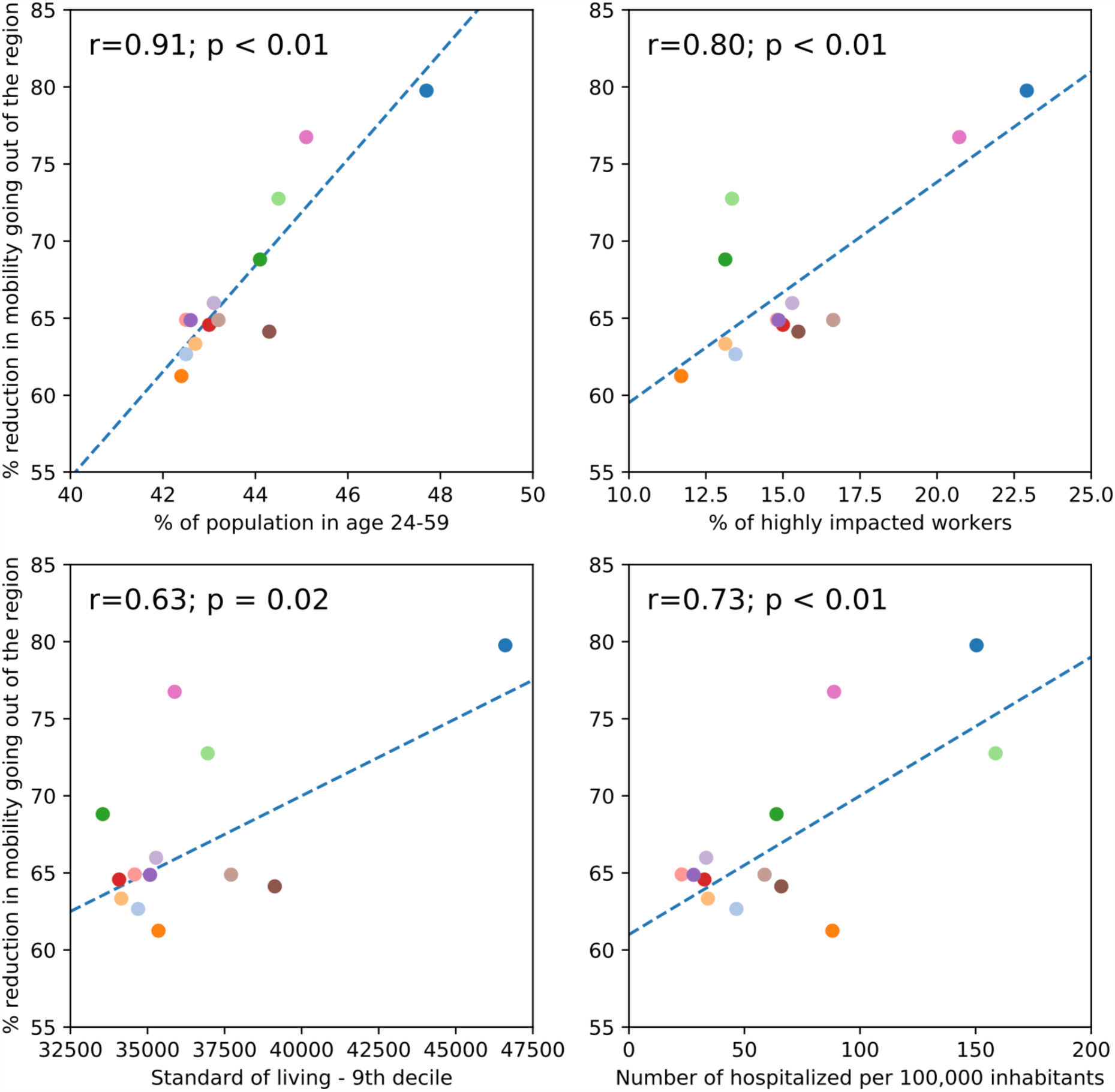
Reduction in outgoing mobility for the week April 6-12, 2020 vs. epidemic, socio-economic, and demographic indicators. Correlation is evaluated between variations in the outgoing traffic and the four considered indicators: a) the population in active age (24-59 years old), b) the fraction of employees in the sectors mostly affected by lockdown. c) the 90^th^ percentile of the regional standard of living^17^, d) the cumulated number of COVID-19 hospitalizations per 100,000 inhabitants on April 05, 2020. Pearson correlation coefficients and their p-values are reported.

Regional drops in mobility in a given week (April 6-12, 2020) were strongly associated with COVID-19 hospitalization rates registered and communicated in the week before (April 5) (Pearson r = 0.73, p < 0.01; **Figure 4**). Analogously, they were strongly correlated to COVID-19-related deaths (Pearson r = 0.63, p = 0.02). Also, hospitalizations and deaths were highly correlated between each other (Pearson r = 0.97, p < 0.01).

Similar results were obtained for drops in mobility within the region, except for the association with the hospitalization rate per region, which however showed a similar, though non-significant, tendency (**Table S1** and **Fig. S1** in **Appendix S4**). Taking out the data point of Île-de-France as the region mostly affected by a departure of inhabitants for relocation in other regions led to similar results (**Table S2** in **Appendix S4**). Appendix S4 also reports a multivariate analysis that includes the behavioral, demographic and socioeconomic factors considered.

### Disruption of mobility connections

We found that some connections completely disappeared, as individuals stopped going from one location to another (**Figure 5**).

**Figure 5.**
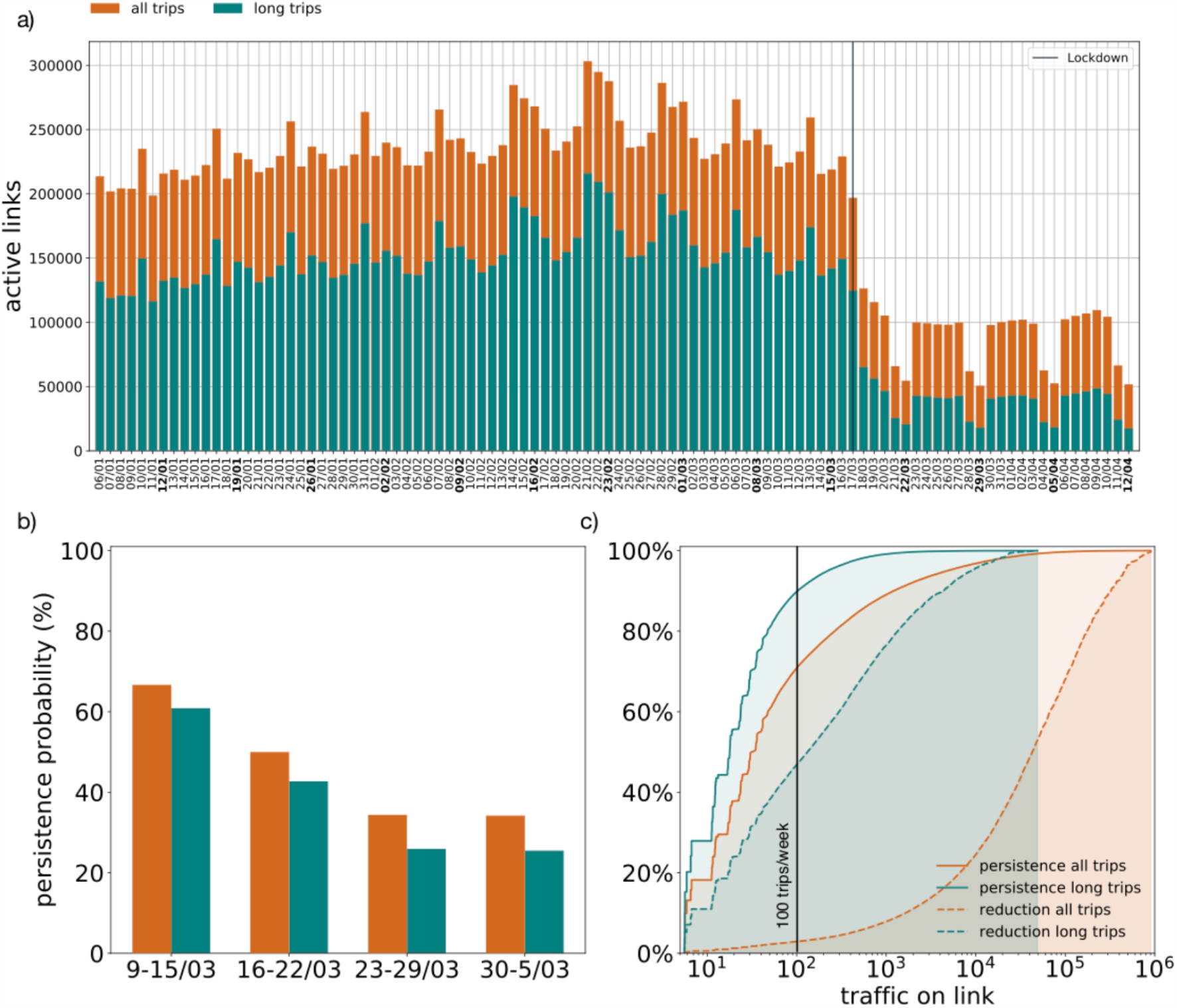
Network analysis. a) Number of mobility connections between French locations over time. b) Link persistence probability: probability that a connection present during week February 3-9 is still present in one of the four selected weeks: before lockdown (March 9-15), during enforcement (March 16-22), during lockdown (March 23-29, March 30 – April 5). c) Persistence probability and traffic reduction in relation with traffic. For a given x-axis value (traffic on link), solid lines measure the fraction of broken links which used to have at most that weight in the baseline week. Dashed lines report the fraction of missing traffic that was lost on connections which used to have at most a certain weight in the baseline week. For all panels: orange: all links, green long links (longer than 100 km).

Connection persistence probability (**Fig. 5b**) decreased steadily during the transition period (67%, 397k over 596k, of connections surviving in the week of school closure and non-essential activity closure announcements, March 9 to 15; 50% in the week of announcement and implementation of lockdown, March 16 to 22). It then stabilized in the first full week of lockdown (34%, 205k over 596k, of connections surviving, March 23) and beyond. Long connections were less resilient than average, as only 1/4 (129k over 497k) of them survived lockdown.

After lockdown effects stabilized, connections characterized by small traffic prior to restrictions (weak connections) were the most likely to disappear, with 70% of them (417k connections) corresponding to 100 trips per week (**Fig. 5c**). The traffic lost on these connections however barely contributed to total traffic reduction (3% contribution, or 7.2M weekly trips). Restricting the analysis to long mobility connections, the fraction of the weak connections disappearing slightly increased (from 70% to 89%, 442k connections), however with a reduction of 47% (5.7M weekly trips) of the traffic.

The disruption in connections occurred with a certain delay compared to reductions in traffic. For example, on Monday March 16 – the day before lockdown – traffic was reduced by 30% with respect to the previous Monday (51M to 36M trips), but the number of connections went down by 4% only (219k to 210k). One week after, traffic drop was 64% (18M trips) and the drop in the number of connections was 55% (94k).

### Mobility connections of 10 most populated French cities

Restrictions on mobility during lockdown had an uneven impact on the 10 most populated French cities. The circle containing 95% of outgoing traffic from each city decreased after lockdown took effect for all cities, indicating that long-range mobility was disrupted more than short-range one (**Figure 6**). But reductions varied from more than 80% (Paris, from 201km to 29km) to 60% (Strasbourg, from 95km to 37km, and Lille, from 78km to 31km), mainly due to different patterns of commuting and connectivity characterizing the mobility of each city.

**Figure 6.**
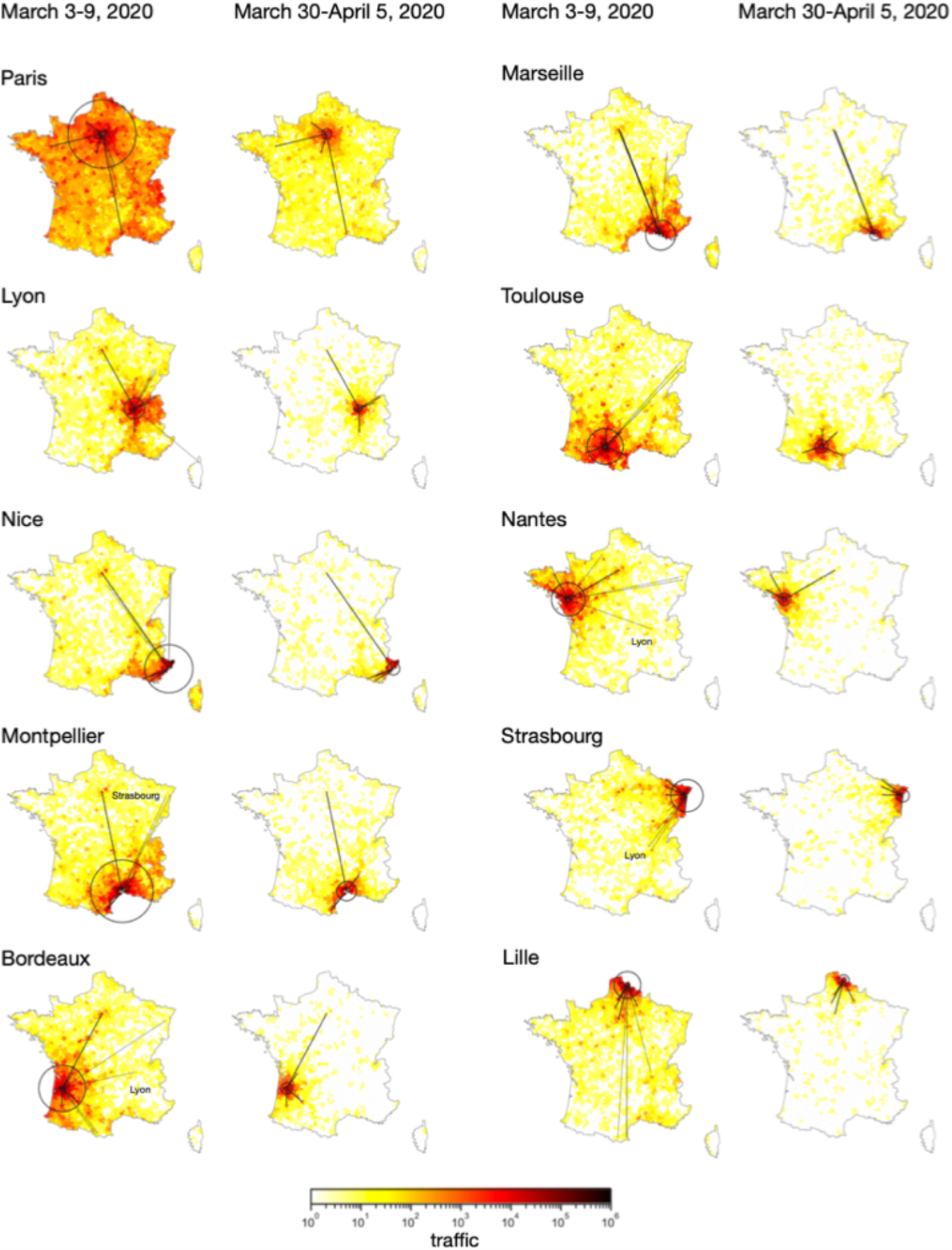
Outgoing egocentric networks of the 10 most populated cities in France during baseline week (starting Feb 3) and during lockdown week starting March 30. Locations are colored by incoming traffic from the selected city. Solid lines indicate links that persist during lockdown. Dashed lines link that disappear. Both are selected to be the top ranking by traffic during the baseline week. The circles contain 95% of the outgoing traffic.

Connections among main cities disappeared too. During lockdown, we no longer detected mobility from Bordeaux, Montpellier, and Nantes to Lyon, or from Montpellier to Strasbourg (**Figure 6**).

## DISCUSSION

Using travel flow data extracted from mobile phone trajectories, we documented a large drop in both short-range and long-range population mobility following lockdown enforcement in France. Overall, trips were reduced by 65%, similarly to reductions found in Belgium^7^, Spain^12^, and Italy^10^ during lockdown, albeit different data sources, spatial resolutions, and definitions of mobility proxies prevent direct numerical comparisons.

The transition signaling the drop in mobility lasted almost a week, anticipating the enforcement of lockdown and creating opposite mobility behaviors. Individuals started spontaneously reducing their mobility on Saturday following the announcement of school closure (March 14), likely because of fear of the growing epidemic and heightened risk aversion^21,22^ generated by the first governmental decision on nationwide interventions. At the same time, fear of an imminent change in policy imposing stricter restrictions – as already implemented in Italy, Spain, Austria - pushed individuals to relocate themselves even to farther away regions where to spend the period of lockdown, if put in place. The exodus, largely covered by the press ^20^, started before the announcement of lockdown and led to anomalous increases in mobility flows out of certain regions (e.g. Île-de-France) and incoming in others (e.g. Normandy). Such behavior was similarly reported in China (out of Wuhan), in Italy (North-to-South), and in India^9^. It demonstrates that the timing at which a policy is announced might disrupt social dynamics as much as the direct effect of the policy, at least in the short term. Increased caution should therefore be considered in the period from announcements to enforcement to avoid unwanted seeding events. These events were not observed in the receiving regions, as lockdown strongly suppressed epidemic activity everywhere. Notwithstanding, they may become important when phasing out restrictions, as less strict social distancing measures may prevent such suppression. Region-specific interventions may increase this risk by inducing similar behavioral responses. New York State reported for instance increased mobility in counties with no imposed lockdown^15^. In this perspective, nationwide interventions and restrictions limiting displacements were adopted by several countries to prevent compensation effects and reduce the possible geographical spread of the epidemic.

Once lockdown entered into effect, population mobility reductions were heterogeneous across regions. Larger reductions were measured in regions more severely hit by the epidemic, with an estimated 1% decrease in regional mobility every 10 additional hospitalizations (per 100,000 inhabitants). A similar association, but with confirmed cases, was observed in Germany ^8^.This suggests that individuals witnessing a larger COVID-19 burden on the hospital system in their region may have further limited displacements compared to those living in less affected regions. Media largely communicated on the epidemic, also providing early on region-specific information on hospitalizations and mounting pressure on the healthcare system. Exposure to this information likely triggered a behavioral response to reduced exposure to the perceived risk, thus increasing compliance to movement restrictions^22^. A similar, though stronger, behavior was observed during a 3-day national lockdown enforced nationwide in Sierra Leone in March 2015 in an effort to control Ebola epidemic^5^. The correlation remains significant even taking out the region of Île-de-France, which experienced a reduction in population due to relocation of individuals. Adjusting for the effect of demography and socioeconomic factors did not qualitatively change the associations found. We also tested COVID-19-related deaths as a proxy for perceived disease severity, and found a similar behavior, explained by the extremely high correlation between hospitalizations and deaths.

Clearly, other factors may have come into play to differentiate drops in regional mobility. Lockdown restrictions had a severe impact on jobs and the organization of work. Regions with the higher proportion of activity sectors mostly impacted by the lockdown (due to telework, but also to complete or partial closure of sectors, such as tourism, entertainment, food services, and construction) also experienced larger drops on mobility. A smaller fraction of active individuals continued to go to work, while the others limited their displacements respecting lockdown mobility restrictions. Indeed, regions with larger portions of the population in the most active age range (24-59y) were also the ones where lockdown had the largest effects. Adjusting for behavioral effects did not change the results.

Uneven mobility drops were also associated with socioeconomic disparities, similarly to what found in Italy^23^. Increasing evidence points at different socioeconomic strata getting uneven shares of the COVID-19 burden^24^. Higher income jobs can often be performed remotely, in confinement, whereas lower income jobs cannot. A survey in France reported that 39% of low income workers were still going to their workplace during lockdown, against only 17% of high income workers^25^. Also, wealthier population strata weather short-term financial losses better, making them more prone to stop working and stay at home if they are afraid or sick^24^. Adjusting for disease perception and demography reversed this association. This is likely due to complex interactions between behavioral response, demography, and economic development. The limited available dataset however does not allow disentangling these effects and infer causal relationships (see Appendix S4).

A strong response was documented in the older age class: as seniors are at highest risk of developing severe forms of COVID-19 if infected, they might also have exhibited increased risk aversion. Specifically, they almost stopped taking trips longer than 100 km, likely to avoid leisure activities and family trips, as recommended by authorities. The most effective reduction in overall mobility occurred during rush hours, associated with a disruption of commuting patterns. This reduction alone likely boosted the role of mobility restrictions in suppressing viral diffusion, as mounting evidence shows that public transportation is a main risk factor for transmission^26^.

Lockdown caused larger disruptions on long-range mobility, as also reported in Belgium^7^ and Italy^10^. Short-range and long-range mobility flows play different roles in the spread of an infectious disease epidemic. Short-range connections are mainly responsible for local diffusion in the community within and around a metropolitan area, whereas long-range connections drive the spatial spread of the epidemic, acting as seeding events to otherwise unaffected or weakly affected areas^27^. Mobility flows out of the city of Wuhan were shown to have seeded other prefectures in China in the early phase of the epidemic before travel restrictions and substantial control measures were implemented^28^. Long-range mobility restrictions therefore contribute to geographically containing the epidemic, especially when epidemic activity shows the patchy geographical pattern observed in many affected countries, including France. Banning trips above 100 km thus helps breaking the spreading pathways, as observed during the lockdown. Nonetheless, we documented that some long-range connections, survived even during lockdown. These movements should be carefully accompanied by strict preventive measures to avoid re-seeding events.

The largest reduction of mobility across distance was reported for Paris. Before lockdown, 95% of outgoing traffic reached destinations within 200 km from the city center, approximately the distance between Paris and Lille, close to the Belgian border. After lockdown, this radius reduced to 29 km, the distance from the city center to Disneyland Paris. Mobility focused around metropolitan areas, serving the needs of individuals in essential professional categories, who continued their work-related displacements. A similar geographical fragmentation was observed in Italy^29^.

Our analysis offered plausible interpretations on how the labor market, demographic and socio-economic indicators, and awareness of increased epidemic risk might have shaped the reduction in mobility, confirming evidence observed in previous^5^ and current^25,30^ outbreaks. Our findings might extend to countries in Europe that qualitatively shared France’s epidemic wave and interventions^8–13^.

Our study has limitations. Despite the widespread use of mobile phone data to quantify mobility^4^, potential sources of inaccuracy traditionally exist: population representativeness, geographic coverage, and heterogeneity in user activity. We used passively-collected signaling data, that improve temporal accuracy and do not depend on activity behavior, compared to traditional call details records. Data owner pre-processed the data to be representative of the general population. Large population displacements may also bias regional activity measures. However, the associations we found were robust after removing Île-de-France, the region mostly affected by the pre-lockdown exodus. Our study is observational, therefore caution is needed in drawing causal relations between the covariates and changes in mobility. Also, the available sample was too small to statistically measure confounding effects rigorously.

Mobility restrictions had the goal of relieving the strain on the healthcare system caused by rapidly increasing hospitalization rates. The effectiveness of these top-down measures was unknown beforehand. Our study showed that different effects were observed across scales, with larger disruptions on long-range connections leading to a localization of the mobility. By associating heterogeneous performances of travel restrictions to both a priori population features (socioeconomic, demographic), and behavioral adaptations to the epidemic and to restrictions themselves, our findings contribute to informing the ongoing epidemic response. Quantitatively characterizing how lockdown reshaped mobility might help design future restrictions that are less pervasive, more targeted.

## Data Availability

Demography, Socio-economic and epidemiological data are available online. Data owners are reported in the main text and the links to download the data are reported in the references. Mobile phone data were provided by the Orange Business Service Flux Vision.

## DATA AVAILABILITY

Mobile phone data are proprietary and confidential. We obtained access to these data from the Orange Business Service Flux Vision within the framework of the research project ANR EVALCOVID-19 (ANR-20-COVI-0007). For legal reasons, the full dataset cannot be published. However, access to the mobility data can be requested from Orange SA on a contractual basis. Hospitalization data were obtained from Santé Publique France and are available at the cited source^16^. Census and socioeconomic data can be accessed through the official website of the French National Statistical Institute (INSEE) at the cited source^17^.

## ACKNOWLEDGMENTS

We thank Luca Ferreri, Sylvain Bourgeois, Erwan Le Quentrec, Zbigniew Smoreda, Chiara Poletto, Pierre-Yves Boëlle for useful discussions.

## AUTHOR CONTRIBUTIONS

EV, SR and VC conceived of and designed the study. GP, EV, and NS analyzed the data, and did the analysis. GP, EV, NS, SR and VC interpreted the results. EV and VC drafted the Article. All authors contributed to the writing of the final version of the article.

## DECLARATIONS OF INTERESTS

GP declares no competing interests. EV declares no competing interests. NS declares no competing interests. SR declares no competing interests. VC declares no competing interests.

## Notes

### Competing Interest Statement

The authors have declared no competing interest.

### Funding Statement

This study is partially funded by: ANR projects EVALCOVID-19 (ANR-20-COVI-0007), SPHINX (ANR-17-CE36-0008-05) and DATAREDUX (ANR-19-CE46-0008-03); EU H2020 grants RECOVER (H2020- 101003589) and MOOD (H2020-874850); REACTing COVID-19 modeling grant.

### Author Declarations

Mobile phone data were previously anonymized in compliance to strict privacy requirements, reviewed and approved by the French National Commission for Data Protection (CNIL, Commission nationale de l'informatique et des libertes), ruling on all matters related to ethics, data, and privacy (https://www.cnil.fr/).

## REFERENCES

1. Anderson, R. M., Heesterbeek, H., Klinkenberg, D. & Hollingsworth, T. D. How will country-based mitigation measures influence the course of the COVID-19 epidemic? The Lancet 395, 931–934 (2020).

2. Di Domenico, L., Pullano, G., Sabbatini, C. E., Boëlle, P.-Y. & Colizza, V. Impact of lockdown on COVID-19 epidemic in Île-de-France and possible exit strategies. BMC Med. 18, 240 (2020).

3. Expected impact of reopening schools after lockdown on COVID-19 epidemic in Île-de-France | medRxiv. https://www.medrxiv.org/content/10.1101/2020.05.08.20095521v1.

4. Lai, S., Farnham, A., Ruktanonchai, N. W. & Tatem, A. J. Measuring mobility, disease connectivity and individual risk: a review of using mobile phone data and mHealth for travel medicine. J. Travel Med. 26, (2019).

5. Peak, C. M. et al. Population mobility reductions associated with travel restrictions during the Ebola epidemic in Sierra Leone: use of mobile phone data. Int. J. Epidemiol. 47, 1562–1570 (2018).

6. Oliver, N. et al. Mobile phone data for informing public health actions across the COVID-19 pandemic life cycle. Sci. Adv. 6, eabc0764 (2020).

7. Covid-19: belgium analyses telecom data to measure the impact of confinement. https://press.telenet.be/covid-19-belgium-analyses-telecom-data-to-measure-the-impact-of-confinement#. Accessed: March 2020.

8. Brockmann, D. et al. Covid-19 Mobility Project in Germany. https://www.covid-19-mobility.org/. Accessed: April 2020.

9. Denis, E., Telle, O. & Benkimoun, S. Mapping the lockdown effects in India: how geographers can contribute to tackle Covid-19 diffusion. The Conversation http://theconversation.com/mapping-the-lockdown-effects-in-india-how-geographers-can-contribute-to-tackle-covid-19-diffusion-136323. Accessed: April 2020.

10. Pepe, E. et al. COVID-19 outbreak response: a first assessment of mobility changes in Italy following national lockdown. medRxiv 2020.03.22.20039933 (2020) doi:10.1101/2020.03.22.20039933.

11. Statista. Community mobility changes due to the coronavirus (COVID-19) outbreak in Poland. https://www.statista.com/statistics/1110080/poland-mobility-changes-due-to-covid-19/. Accessed: July 2020.

12. José Javier Ramasco, M. M. Ramasco, J. J., and Mazzoli, M., 4° Informe sobre los cambios de movilidad en España debido a las medidas de confinamiento contra la extensión del COVID-19 https://analytics.ifisc.uib-csic.es/es/respuesta-covid-19/?fbclid=IwAR2C8nA_CWa_AgrD4e_DF5UqWVs-QNZpQeDkxGED-HS1gOyDX5aDpr6UZKY Accessed?: April 2020.

13. Santana, C., Botta, F., Barbosa, H., Privitera, F. & Clemente, R. D. Analysis of human mobility in the UK during the COVID-19 pandemic. doi:10.13140/RG.2.2.33207.14240.

14. Bakker, M., Berke, A., Groh, M., Pentland, S. & Moro, E. Effect of social distancing measures in the New York City metropolitan area. http://curveflattening.media.mit.edu/Social_Distancing_New_York_City.pdf. Accessed: April 2020.

15. Gao, S., Rao, J., Kang, Y., Liang, Y. & Kruse, J. Mapping County-Level Mobility Pattern Changes in the United States in Response to COVID-19. https://papers.ssrn.com/abstract=3570145 (2020) doi:10.2139/ssrn.3570145.

16. santepubliquefrance.fr. Indicateurs?: cartes, données et graphiques. https://geodes.santepubliquefrance.fr/#c=indicator&f=0&i=covid_hospit.hosp&s=2020-04-09&t=a01&view=map1. Accessed: April 2020.

17. INSEE. https://www.insee.fr. Accessed: April 2020.

18. DARES. Activité et conditions d’emploi de la main-d’œuvre pendant la crise sanitaire Covid-19. https://dares.travail-emploi.gouv.fr/IMG/pdf/dares_acemo_covid19_synthese_17-04-2020.pdf. Accessed: May 2020.

19. Prophet. https://facebook.github.io/prophet/. Accessed: April 2020.

20. Le Figaro. Coronavirus?: le grand exode des citadins. https://www.lefigaro.fr/secteur/high-tech/le-grand-exode-des-citadins-20200327. Accessed: March 2020.

21. Rubin, G. J., Amlôt, R., Page, L. & Wessely, S. Public perceptions, anxiety, and behaviour change in relation to the swine flu outbreak: cross sectional telephone survey. BMJ 339, (2009).

22. Funk, S., Salathé, M. & Jansen, V. A. A. Modelling the influence of human behaviour on the spread of infectious diseases: a review. J. R. Soc. Interface 7, 1247–1256 (2010).

23. Bonaccorsi, G. et al. Economic and Social Consequences of Human Mobility Restrictions Under COVID-19. https://papers.ssrn.com/abstract=3573609 (2020) doi:10.2139/ssrn.3573609.

24. The Economist. American inequality meets covid-19. https://www.economist.com/united-states/2020/04/18/american-inequality-meets-covid-19. Accessed: April 2020.

25. Chloé Morin, Jérôme Fourquet, Marie Le Vern. Premiers de corvée et premiers de cordée, quel avenir pour le travail déconfiné?. https://jean-jaures.org/nos-productions/premiers-de-corvee-et-premiers-de-cordee-quel-avenir-pour-le-travail-deconfine. Accessed: April 2020.

26. Bi, Q. et al. Epidemiology and transmission of COVID-19 in 391 cases and 1286 of their close contacts in Shenzhen, China: a retrospective cohort study. Lancet Infect. Dis. 20, 911–919 (2020).

27. Balcan, D. et al. Multiscale mobility networks and the spatial spreading of infectious diseases. Proc. Natl. Acad. Sci. 106, 21484–21489 (2009).

28. Jia, J. et al. Population flow drives spatio-temporal distribution of COVID-19 in China. Nature 1–11 (2020) doi:10.1038/s41586-020-2284-y.

29. Bonato, P. et al. Mobile phone data analytics against the COVID-19 epidemics in Italy: flow diversity and local job markets during the national lockdown. arXiv 2004.11278, (2020).

30. The New York Times. Location Data Says It All: Staying at Home During Coronavirus Is a Luxury. https://www.nytimes.com/interactive/2020/04/03/us/coronavirus-stay-home-rich-poor.html. Accessed: April 2020.

